# Differentials in the characteristics of COVID-19 cases in Wave-1 and Wave-2 admitted to a network of hospitals in North India

**DOI:** 10.1101/2021.06.24.21259438

**Authors:** Sandeep Budhiraja, Abhaya Indrayan, Mona Aggarwal, Vinita Jha, Dinesh Jain, Bansidhar Tarai, Poonam Das, Bharat Aggarwal, RS Mishra, Supriya Bali, Monica Mahajan, Vivek Nangia, Ajay Lall, Nevin Kishore, Ashish Jain, Omender Singh, Namrita Singh, Ashok Kumar, Prashant Saxena, Arun Dewan, Ritesh Aggarwal, Shailesh Sahay, Rajiv Dang, Neelima Mishra, Mohit Mathur, I. M. Chugh, Pankaj Aneja, Sanjay Dhall, Vandana Boobna, Vinit Arora, Ajay Gupta, Vijay Arora, Mukesh Mehra, Meenakshi Jain, Vimal Nakra, BD Sharma, Praveen Pandey, YP Singh, Anil Vardani, RK Singal, Deepak Gargi Pandey, Atul Bhasin, Sandeep Nayyar, Rajesh Pande, Pankaj Chaudhary, Ajay Kr Gupta, Ashish Gupta, Nitesh Tayal, Puneet Gupta, Manish Gupta, Sumit Khetrapal, Sachin Pandove, Deepak Bhasin, Devender Midha, Harpal Singh, Ambrish Dixit, Vinay Sagar, Vaibhav Chachra, Bhupesh Uniyal, Sanjay Saxena, Amarjit Singh, Shalini Sharma

**Affiliations:** Max Super Speciality Hospital, Saket, New Delhi, India; Department of Clinical Directorate, Max Healthcare, New-Delhi, India; Department of Laboratory Sciences, Max Super Speciality Hospital, Saket, New Delhi, India; Department of Radiology, Max Super Speciality Hospital, Saket, New Delhi, India; Max Smart Super Speciality Hospital, Saket, New Delhi, India; Max Hospital, Gurgaon, Haryana; Max Super Speciality Hospital, Shalimar Bagh, New Delhi, India; Max Super Speciality Hospital, Patparganj, New Delhi, India; BLK-Max Super Speciality Hospital, New Delhi, India; Max Super Speciality Hospital, Vaishali, U.P. India; Max Super Speciality Hospital, Mohali, Punjab, India; Max Super Speciality Hospital, Dehradun, Uttarakhand, India; Max Super Speciality Hospital, Bhatinda, Punjab, India

**Keywords:** COVID-19, Cases in Wave-1 and Wave-2, Demography, Mortality, Wave differentials

## Abstract

Second wave of COVID-19 pandemic in India came with unexpected quick speed and intensity, creating an acute shortage of beds, ventilators, and oxygen at the peak of occurrence. This may have been partly caused by emergence of new variant delta. Clinical experience with the cases admitted to hospitals suggested that it is not merely a steep rise in cases but also possibly the case-profile is different. This study was taken up to investigate the differentials in the characteristics of the cases admitted in the second wave versus those admitted in the first wave.

Records of a total of 14398 cases admitted in the first wave (2020) to our network of hospitals in north India and 5454 cases admitted in the second wave (2021) were retrieved, making it the largest study of this kind in India. Their demographic profile, clinical features, management, and outcome was studied.

Age-sex distribution of the cases in the second wave was not much different from those admitted in the first wave but the patients with comorbidities and those with greater severity had larger share. Level of inflammatory markers was more adverse. More patients needed oxygen and invasive ventilation. ICU admission rate remained nearly the same. On the positive side, readmissions were lower, and the duration of hospitalization was slightly less. Usage of drugs like remdesivir and IVIG was higher while that of favipiravir and tocilizumab was lower. Steroid and anticoagulant use remained high and almost same during the two waves. More patients had secondary bacterial and fungal infections in Wave-2. Mortality increased by almost 40% in Wave-2, particularly in the younger patients of age less than 45 years. Higher mortality was observed in those admitted in wards, ICU, with or without ventilator support and those who received convalescent plasma.

No significant demographic differences in the cases in these two waves, indicates the role of other factors such as delta variant and late admissions in higher severity and more deaths. Comorbidity and higher secondary bacterial and fungal infections may have contributed to increased mortality.

## INTRODUCTION

Many countries reported two-wave pattern of COVID-19 since the start of pandemic in early 2020. A large majority of these countries were relatively unprepared during the first wave. Due to factors such as high sero-prevalence amongst dense population clusters, better infrastructure, more robust management guidelines, accessible, faster and cheaper diagnostic tests, and, most importantly, availability of vaccines after the first wave, likelihood of an explosive second wave was being considered less likely. But several countries experienced a catastrophic second wave, and saw the contagion to be much more infectious and, in some places, probably more virulent. Many countries started reporting increasing occurrence of variants of concern (VOC) like the UK strain (B.1.17), South African strain (B.1.351), Brazilian strain (P.1) and finally the Indian strain (B.1.617.2). Recently, the nomenclature of these has been changed by WHO to alpha (UK strain; B.1.17), beta (South African strain; B.1.351), gamma (Brazilian strain; P.1) and delta (Indian strain; B.1.617.2) strains. [1]

Many countries reported steep second wave due to these VOC [2, 3, 4]. In UK, the second wave starting January 2021 was largely due to the alpha variant. India also reported increasing alpha variant in early 2021 but this soon got replaced by the delta strain that led to the explosion of cases in April and May 2021, leading to a huge second wave [2, 4, 5]. The cases of break-through infection following vaccination have also been reported from many areas. [4, 5, 6].

The second wave silently began in India in January 2021 and it became steep in April, attaining peak in early May. The surge of cases was so sudden that everybody was taken unawares, and the news of huge shortage of beds, oxygen, and ventilators emerged. This may have caused unspecified number of avoidable misery and deaths. We observed for the cases admitted to our network of hospitals that it is not simply the steep rise in cases, but they now have a very different clinical and laboratory profile. This could have been due to wide spread affliction by the new variant of concern, namely the B.1.167.2 [3, 6].

The present study was undertaken to investigate differentials in the cases admitted in the second wave compared to the first wave. Besides documentation, this may shed light on the cause and the characteristics of the cases in second wave.

## METHODS

Ten hospitals, spread across five north Indian states, using the same Electronic Health Record (EHR), that have been admitting and treating COVID-19 patients since April 2020, were included in this retrospective study. All consecutive RT-PCR positive COVID-19 patients admitted to these hospitals and for whom EHR data was accessible were included. Wave-1 was defined as period from April to December 2020 and Wave-2 from January to June 2021 for the purpose of this study.

The number of weekly cases admitted to ICU, ward and total, in our network of hospitals followed the same peak (Figure 1) as in the general population and both the waves corresponded fairly well to the RT-PCR positivity among the tests done in our laboratories (Figure 2).

**Figure 1:**
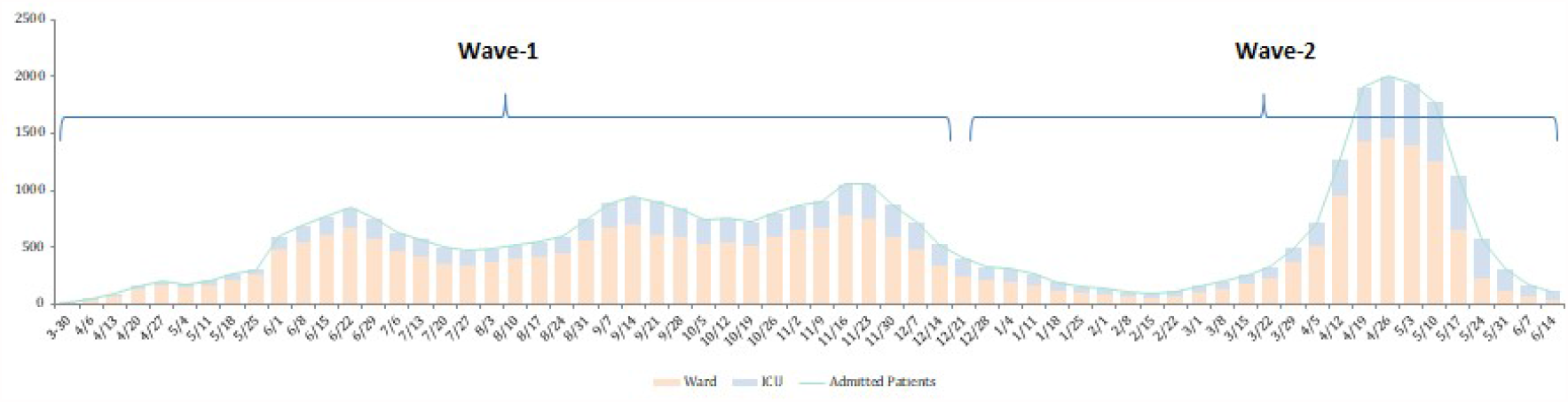
Average number of COVID-19 patients admitted in our network (weekly)

**Figure 2:**
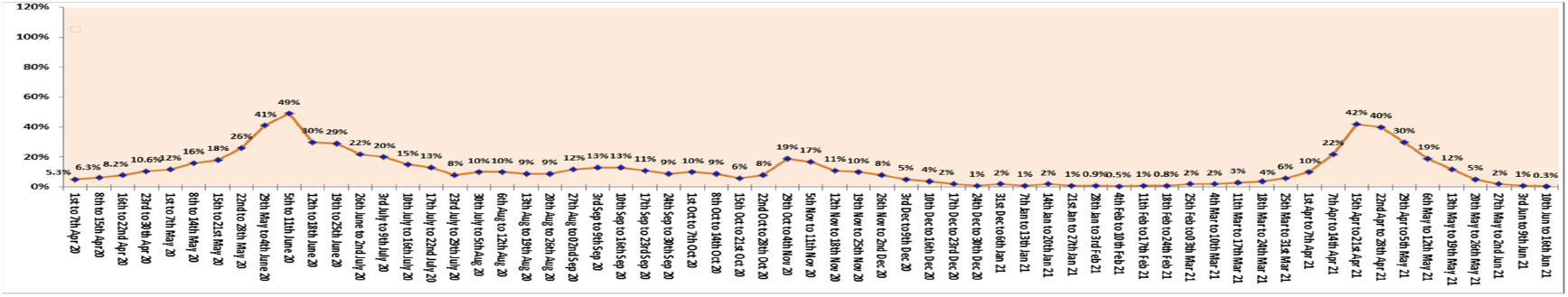
RT-PCR positivity percentage in Wave-1 and Wave-2

The characteristics under comparison included severity of COVID-19 at admission, demographics (age and sex), co-morbidity, admission to ward or ICU, duration of hospitalization, oxygen and ventilator support, the drugs used for treatment, laboratory parameters, CT severity score, secondary infections, and mortality. Many parameters that were compared in this study were not available for the entire cohort and hence various subsets had differing sizes.

The severity of COVID-19 was classified as mild, moderate, and severe, as per the criteria of Ministry of Health and Family Welfare [7].

All the admitted patients gave a prior consent for their anonymised data to be used for research purpose.

### Ethics Committee Approval

This study was approved by the Institutional Ethics Committee, Max Super Speciality Hospital (A unit of Devki Devi Foundation), address : Service Floor, Office of Ethics Committee, East Block, next to Conference Room, Max Super Speciality Hospital, Saket (A unit of Devki Devi Foundation), 2, Press Enclave Road, Saket, New Delhi – 110017 vide ref. no. BHR/RS/MSSH/DDF/SKT-2/IEC/IM/21-15 dated 23^rd^ June’2021. The IEC provided no objection and approval for the publication of above manuscript.

### Statistical Methods

The percentage of cases with different characteristics in the two waves was compared by chi-square test. The laboratory parameters had highly skewed distribution and they were collated with median and inter-quartile range (IQR) and compared with nonparametric Mann-Whitney test. A P-value less than 0.05 was considered statistically significant although, in this case, the number of cases is so large for some categories that P-values have to be cautiously interpreted. SPSS21 was used for calculations.

## RESULTS

A total of 19,852 RT-PCR confirmed COVID-19 patients were included in the present study. During the period from April to December 2020, 14398 patients were included (Wave-1) and majority of admissions took place in the months of June, September and November 2020. For the period January to mid June 2021, 5454 patients were included (Wave-2) and majority of admission happened in April and May 2021.

### Background Characteristics

Among the cases admitted to our hospitals, mild cases were relatively lower (P<0.001) and severe cases higher (P < 0.001) in the second wave (Table 1).

**Table 1.** Severity of cases admitted in Wave-1 and Wave-2

| Severity | Wave-1 (n = 14398) | Wave-2 (n = 5454) | P-value |
| --- | --- | --- | --- |
| Mild | 4986 (34.6%) | 1416 (26.0%) | <0.001 |
| Moderate | 4707 (32.7%) | 1891 (34.7%) | 0.008 |
| Severe | 4705 (32.7%) | 2147 (39.4%) | <0.001 |

Overall, in each wave, almost two-thirds were males. Females were admitted slightly more in Wave-2 compared to the Wave-1 (36.3% vs 32.6%, P<0.001). Age distribution remained nearly same in both the waves (P = 0.143), and the age group 60+ years continued to have disproportionately large share (nearly 40%). Relative to their population (less than 10% at the all-India level in this age-group), this age group was nearly four times as likely to be admitted. Patients of age less than 45 years comprised 28.3% and 27.1% in Wave-1 and Wave-2, respectively (Table 2). Comorbidities were more commonly present (P<0.001) in patients in Wave-2 (59.7%) than in Wave-1 (54.8%). In particular, during Wave-2, diabetes (P=0.031), hypertension (P=0.001), and chronic kidney disease (P=0.004) were significantly more common but not coronary artery disease (P=0.106).

**Table 2.** Demographic characteristics of the patients in Wave-1 and Wave-2

| Characteristic |  | Wave-1 |  | Wave-2 |  | P-value |
| --- | --- | --- | --- | --- | --- | --- |
|  |  | Number | Percent | Number | Percent |  |
| Total Cases |  | 14398 | 100 | 5454 | 100 |  |
| Sex | Male | 9703 | 67.4 | 3472 | 63.7 | <0.001 |
|  | Female | 4695 | 32.6 | 1982 | 36.3 |  |
| Age | <45 Years | 4069 | 28.3 | 1478 | 27.1 | 0.143 |
|  | 45-59 Years | 4605 | 32.0 | 1764 | 32.3 |  |
|  | 60-74 Years | 4436 | 30.8 | 1676 | 30.7 |  |
|  | ≥75 Years | 1288 | 8.9 | 536 | 9.8 |  |
| Comorbidity | Comorbidity | 7890 | 54.8 | 3256 | 59.7 | <0.001 |
|  | DM | 6220 | 43.2 | 2449 | 44.9 | 0.031 |
|  | HTN | 5903 | 41.0 | 2383 | 43.7 | 0.001 |
|  | CKD | 1958 | 13.6 | 829 | 15.2 | 0.004 |
|  | CAD | 806 | 5.6 | 338 | 6.2 | 0.106 |

### Clinical Characteristics

Nearly one-third of the patients needed ICU/HDU admission, and this proportion remained nearly the same in both the waves (34.9% in Wave-1 vs 33.4% in Wave-2). Readmission were less in the second wave (10% in Wave-2 compared to 14.6% in Wave-1). The average duration of hospitalization was lesser in Wave-2 compared to Wave-1 (Table 3). More than one-fifth (21.4%) patients stayed for less than 5 days in the hospital in Wave-2 compared to 15.7% in Wave-1. Cases with long stay (≥15 days) reduced from 10.4% in Wave-1 to 7% in Wave-2.

**Table 3.**
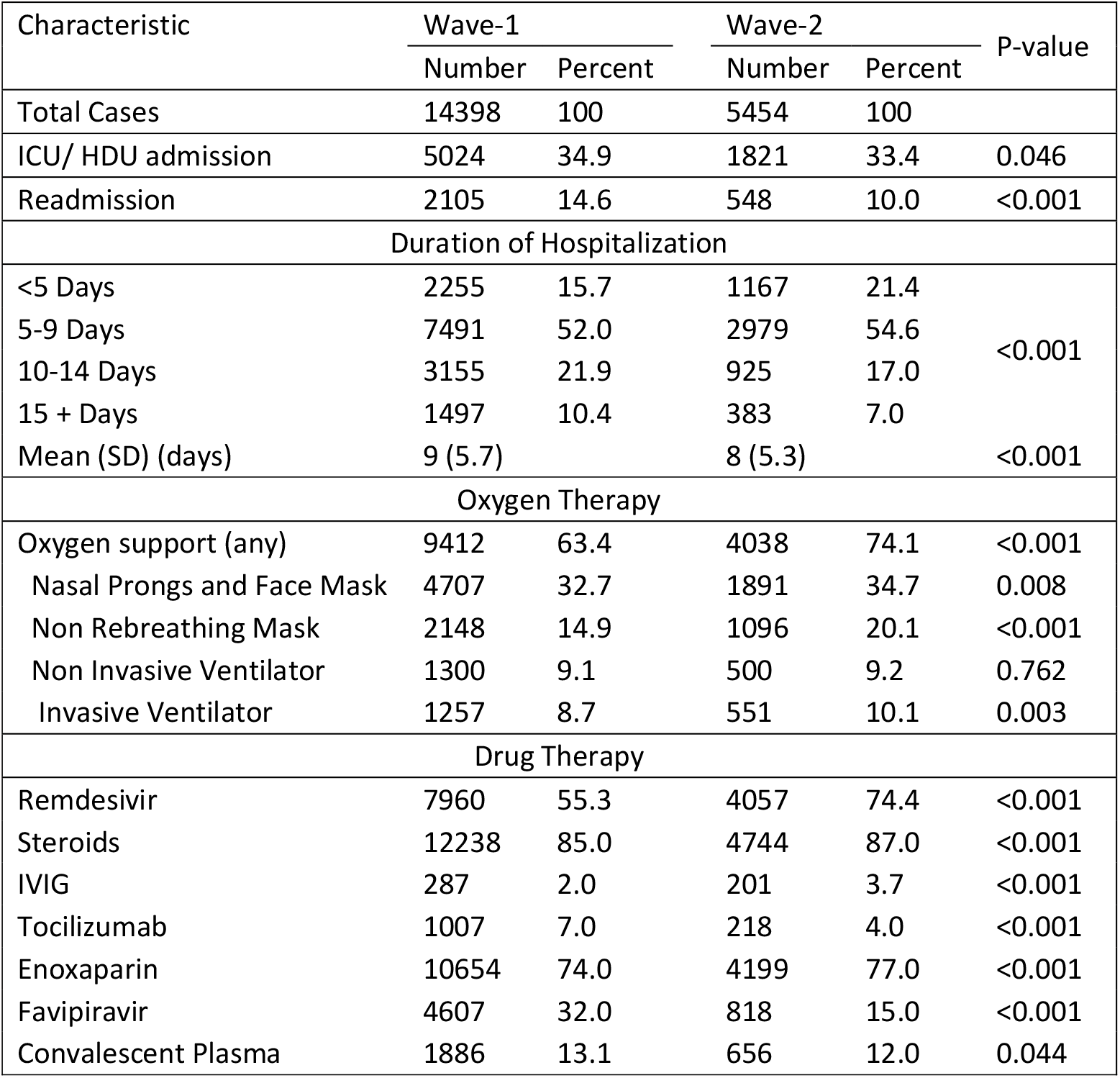
Clinical characteristics of the patients admitted in Wave-1 and Wave-2

| Characteristic | Wave-1 |  | Wave-2 |  | P-value |
| --- | --- | --- | --- | --- | --- |
|  | Number | Percent | Number | Percent |  |
| Total Cases | 14398 | 100 | 5454 | 100 |  |
| ICU/ HDU admission | 5024 | 34.9 | 1821 | 33.4 | 0.046 |
| Readmission | 2105 | 14.6 | 548 | 10.0 | <0.001 |
| Duration of Hospitalization |  |  |  |  |  |
| <5 Days | 2255 | 15.7 | 1167 | 21.4 | <0.001 |
| 5-9 Days | 7491 | 52.0 | 2979 | 54.6 |  |
| 10-14 Days | 3155 | 21.9 | 925 | 17.0 |  |
| 15 + Days | 1497 | 10.4 | 383 | 7.0 |  |
| Mean (SD) (days) | 9 (5.7) |  | 8 (5.3) |  | <0.001 |
| Oxygen Therapy |  |  |  |  |  |
| Oxygen support (any) | 9412 | 63.4 | 4038 | 74.1 | <0.001 |
| Nasal Prongs and Face Mask | 4707 | 32.7 | 1891 | 34.7 | 0.008 |
| Non Rebreathing Mask | 2148 | 14.9 | 1096 | 20.1 | <0.001 |
| Non Invasive Ventilator | 1300 | 9.1 | 500 | 9.2 | 0.762 |
| Invasive Ventilator | 1257 | 8.7 | 551 | 10.1 | 0.003 |
| Drug Therapy |  |  |  |  |  |
| Remdesivir | 7960 | 55.3 | 4057 | 74.4 | <0.001 |
| Steroids | 12238 | 85.0 | 4744 | 87.0 | <0.001 |
| IVIG | 287 | 2.0 | 201 | 3.7 | <0.001 |
| Tocilizumab | 1007 | 7.0 | 218 | 4.0 | <0.001 |
| Enoxaparin | 10654 | 74.0 | 4199 | 77.0 | <0.001 |
| Favipiravir | 4607 | 32.0 | 818 | 15.0 | <0.001 |
| Convalescent Plasma | 1886 | 13.1 | 656 | 12.0 | 0.044 |

More patients required oxygen support this time (74.1% vs 63.4% earlier, P<0.001) and similarly a higher number required invasive ventilation (10.1% in Wave-2 vs 8.7% in Wave-1, P=0.002).

We tried to analyse the use of various treatment options during the two waves. Remdesivir use increased significantly in Wave-2 (74.4% vs 55.3% last year) but usage of steroids remained almost same but high (nearly 86%) and so also of anticoagulants (Enoxaparin) (74-77%). Usage of Intravenous immunoglobulins (IVIG) in Wave-2 was much higher than in Wave-1 (3.7% vs 2%) while that of favipiravir (32% in Wave-1 versus 15% in Wave-2) and tocilizumab (7% in Wave-1 versus 4% in Wave-2) reduced to almost half during Wave-2. Usage of convalescent plasma (CP) remained almost same, 13.1% in Wave-1 vs 12% in Wave-2. (Table 3).

### Laboratory Parameters

The laboratory parameter investigated are Absolute lymphocyte count (ALC), C-reactive protein (CRP), Interleukin-6 (IL-6), D-Dimer, Lactate dehydrogenase (LDH), Ferritin, Creatinine phosphokinase (CPK), and Troponin-I. The results of the comparison of their values at the time of admission in Wave-1 and Wave-2 are in Table 4.

**Table 4.** Median values of various laboratory parameters at the time of admission in Wave-1 and Wave-2

| Laboratory parameter | Wave-1 |  |  | Wave-2 |  |  | P-value comparing 2 waves |
| --- | --- | --- | --- | --- | --- | --- | --- |
|  | n | Median | IQR | n | Median | IQR |  |
| ALC ($10^9$/L) | 12069 | 1.21 | 0.79-1.75 | 4551 | 1.03 | 0.7-1.5 | <0.001 |
| CRP (mg/dl) | 9891 | 8.6 | 1.9-31.7 | 3779 | 13.8 | 3.4-55.5 | <0.001 |
| IL6 (pg/mL) | 8190 | 21.7 | 7.6-57.7 | 3047 | 21.2 | 7.8-54.6 | 0.397 |
| D Dimer (ng/ml) | 9789 | 231.4 | 141-440 | 3701 | 234.1 | 158-411 | <0.001 |
| LDH (U/L) | 7203 | 300 | 234-397 | 2570 | 305.1 | 235-418 | 0.021 |
| Ferritin (ng/mL) | 8821 | 233.8 | 101-487 | 3074 | 244.3 | 106-510 | 0.084 |
| CPK (U/L) | 4389 | 97 | 58-193 | 1742 | 108 | 62-220 | <0.001 |
| Trop I (ng/ml) | 4212 | 0 | 0.01-0.02 | 1777 | 0 | 0.01-0.02 | 0.029 |

Median values of ALC were significantly lower and those of CRP, D-Dimer, LDH and CPK were significantly higher in Wave-2 than in Wave-1. CRP was particularly high in Wave-2 cases – reaching to about one-and-a-half times in the second wave.

### Opportunistic Infections

This analysis was carried out for cases admitted to only one hospital where the records were complete. The number of admissions in this hospital in Wave-1 (May-October 2020) was 3691 and in Wave-2 (March-June 2021) was 1264. Secondary infections were categorised as: blood stream infection (BSI), hospital acquired pneumonia (HAP), urinary tract infections (UTI), and skin and soft tissue infections (SSTI). Opportunistic infections occurred in 439 of 3691 (11.9%) patients in Wave-1 for which the data was available versus 350 out of 1264 (27.8%) during Wave-2 (P<0.001) (Figure 3a). From those who had secondary infections, percentage of patients reporting UTI reduced from 73% in Wave-1 to 38% in Wave-2 though it constituted the most common type of opportunistic infection in both the waves. The percentage of BSI patients increased from 11.0% in Wave-1 to 28.0% in Wave-2, those with HAP increased from 14.0% to 25.0%, and SSTI increased from 3.0% to 10.0%(Figure 3 b, 3 c).

**Figure 3:**
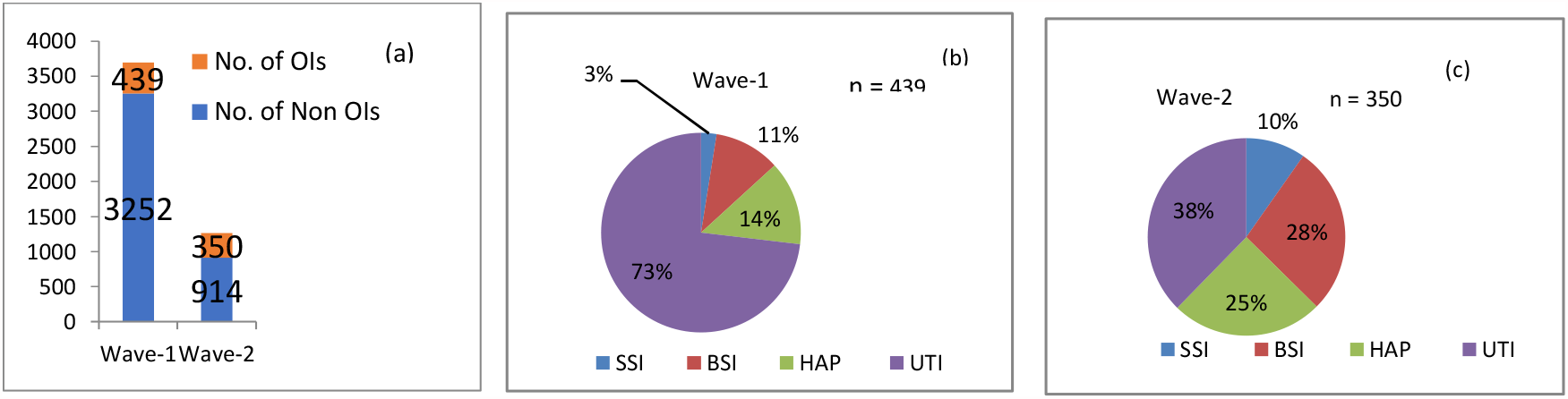
Opportunistic infections (OIs) in hospitalized COVID-19 cases during Wave-1 and Wave-2

In the cases of SSTI, the most common organisms during Wave-1 were Pseudomonas and Klebsiella, while in Wave-2 were E.coli and Staphylococcus (Figure 4a). For BSI, the commonest organisms in Wave-1 were Candida auris (28%), Klebsiella and E coli. In Wave-2, there was no reported case of Candida auris in BSI and common organisms were Klebsiella, E coli & Acinetobacter (Figure 4b). In cases of HAP, during Wave-1, Klebsiella and Pseudomonas were the most commonly isolated organisms while in Wave-2, it was Klebsiella and Acinetobacter (Figure 4c). For UTI, in Wave-1, the common bugs were E coli and Klebsiella. During Wave-2, other than these two, we also identified Enterococcus faecium and Candida tropicalis, in significant proportions (Figure 4d). Figure 5 depicts the reporting of Mucormycosis cases, with its associated mortality, in our hospitals during the two waves. Most of the cases presented after an average of 3 weeks from the disease onset and majority were referred for treatment of Mucormycosis from other non-network hospitals. COVID-associated mucormycosis (CAM) presented as another epidemic in India during the months of May and June 2021. While only 10 cases were admitted to our network of hospitals during Wave-1, with 2 deaths; this number increased to 169 cases with 17 deaths, during Wave-2 (May-June 2021) (Figure 5).

**Figure 4(a):**
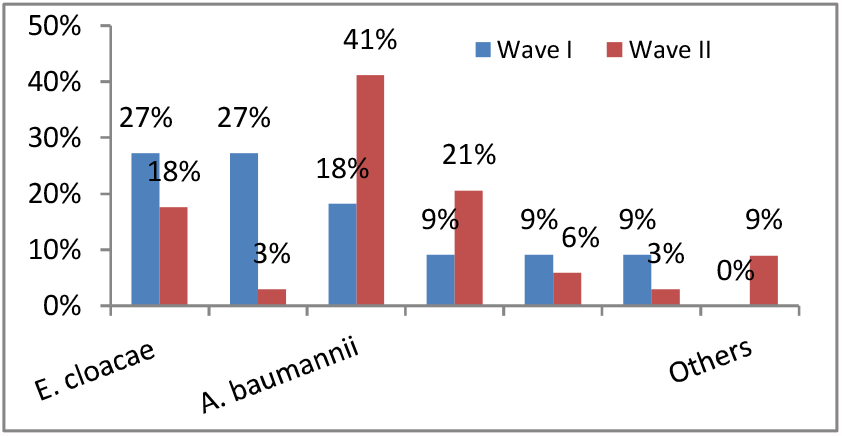
Skin and soft tissue infections in Wave-1 and Wave-2

**Figure 4(b):**
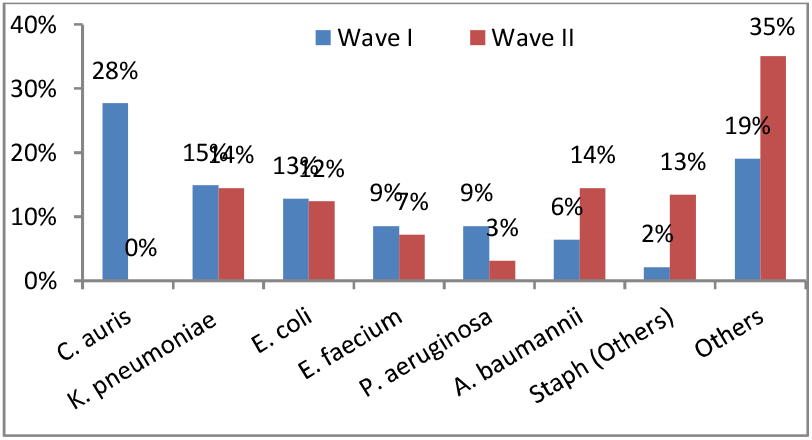
Blood stream infections in Wave-1 and Wave-2

**Figure 4(c):**
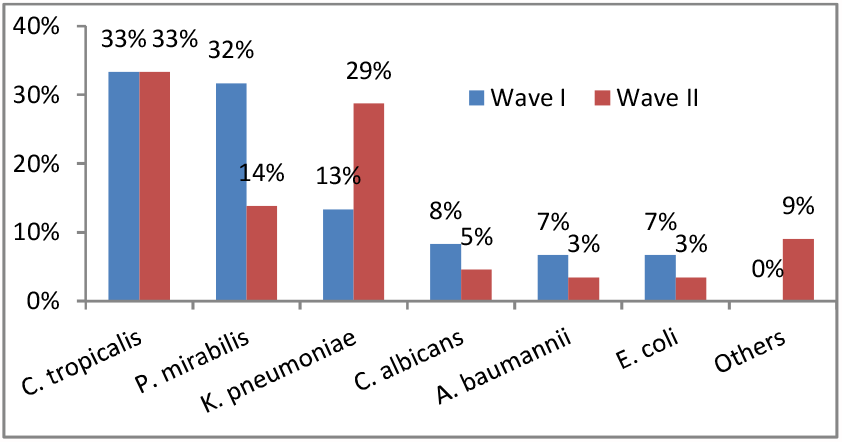
Hospital acquired pneumonia in Wave-1 and Wave-2

**Figure 4(d):**
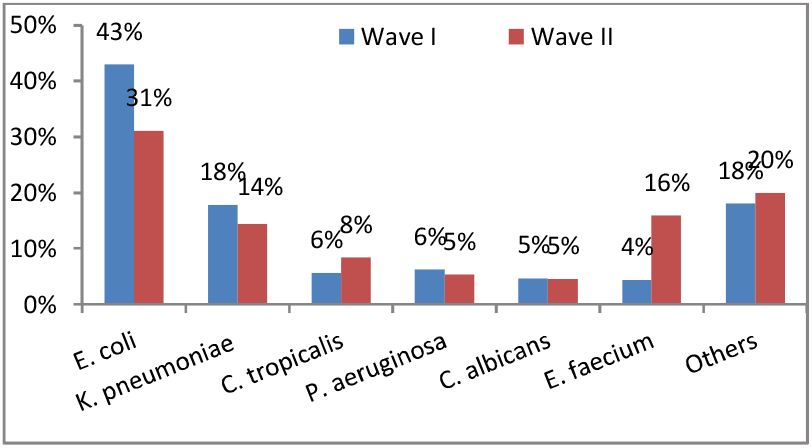
Urinary tract infections in Wave-1 and Wave-2

**Figure 5:**
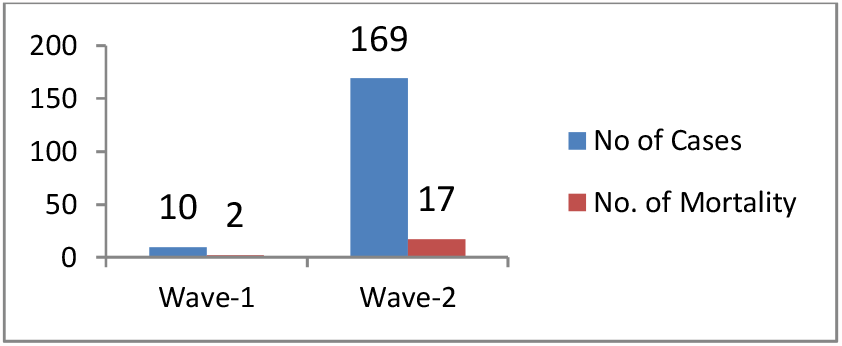
Mucormycosis Cases and deaths in Wave-1 and Wave-

### Mortality

Overall mortality in Wave-2 was nearly 40% higher than in Wave-1 (10.5% vs. 7.2%, P<0.001). This increase in mortality was seen in both males (10.8% vs. 7.4%, P<0.001) and females (9.8% vs. 6.8%, P<0.001). Younger patients (<45 years) saw the sharpest increase in mortality to 4.1% from 1.3% in Wave-1. In this group, it was observed that average duration of symptoms prior to admission was 7.3 days in Wave-2 versus 6.3 days in Wave-1. In this age group, mortality increased in both males (4.7% in Wave-2 vs. 1.4% in Wave-1) and females (2.8% vs. 1.0%). Sex differentials in other age group were not significant. The increasing trend in mortality (Wave-1 vs. Wave-2) was seen across all other age groups also: 45-59 years (5% vs 7.6%), 60-74 years (12% vs. 13.8%) and in ≥ 75 years (18.9% vs. 26.9%) (Figure 6). The higher mortality rates during Wave-2 were seen across various treatment modalities, whether the patients were on non-invasive ventilator (NIV) (40.8% in Wave-1 vs. 48.4% in Wave-2), on invasive ventilator (62.5% vs. 68.4%) and even for those who were not on any ventilator support and those who received convalescent plasma (21.3% vs. 27.6%). Not only was the mortality higher in Wave-2 for patients in ICU (19.8% vs. 25.1%) but steeply higher for those admitted in wards (0.5% vs. 3.1%). (Table 5.)

**Figure 6.**
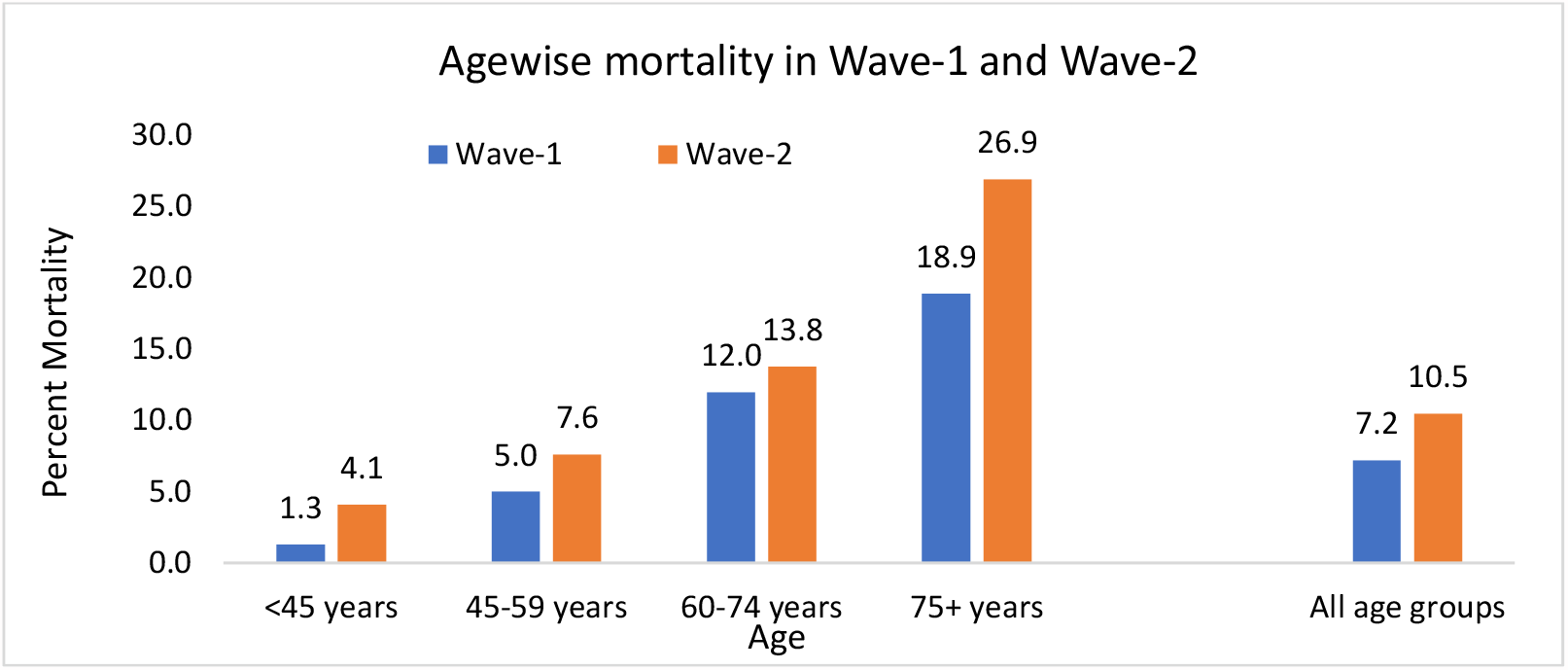
Age wise mortality in Wave-1 and Wave-2

**Table 5.** Mortality in Wave-1 and Wave-2

| Characteristic |  | Wave-1 |  |  | Wave-2 |  |  | P-value for difference between Wave-1 and Wave-2 |
| --- | --- | --- | --- | --- | --- | --- | --- | --- |
|  |  | Cases | Deaths | Percent Mortality | Cases | Deaths | Percent Mortality |  |
| Total Cases |  | 14398 | 1039 | 7.2 | 5454 | 570 | 10.5 | <0.001 |
| Sex | Male | 9703 | 721 | 7.4 | 3472 | 375 | 10.8 | <0.001 |
|  | Female | 4695 | 318 | 6.8 | 1982 | 195 | 9.8 | <0.001 |
| Age | <45 Years | 4069 | 51 | 1.3 | 1478 | 60 | 4.1 | <0.001 |
|  | Male | 2723 | 38 | 1.4 | 984 | 46 | 4.7 | <0.001 |
|  | Female | 1346 | 13 | 1 | 494 | 14 | 2.8 | 0.005 |
|  | 45-59 Years | 4605 | 231 | 5 | 1764 | 134 | 7.6 | <0.001 |
|  | 60-74 Years | 4436 | 531 | 12 | 1676 | 232 | 13.8 | 0.009 |
|  | >75 Years | 1288 | 244 | 18.9 | 536 | 144 | 26.9 | <0.001 |
| On Non Invasive Ventilator | Yes | 1300 | 531 | 40.8 | 500 | 242 | 48.4 | 0.002 |
|  | No | 13098 | 508 | 3.9 | 4954 | 328 | 6.6 | <0.001 |
| On Invasive Ventilator | Yes | 1257 | 785 | 62.5 | 551 | 377 | 68.4 | 0.008 |
|  | No | 13141 | 254 | 1.9 | 4903 | 193 | 3.9 | <0.001 |
| Convalescent Plasma | Yes | 1886 | 402 | 21.3 | 656 | 181 | 27.6 | 0.001 |
|  | No | 12512 | 637 | 5.1 | 4798 | 389 | 8.1 | <0.001 |
| Admission | Ward | 9374 | 43 | 0.5 | 3633 | 113 | 3.1 | <0.001 |
|  | ICU / HDU | 5024 | 996 | 19.8 | 1821 | 457 | 25.1 | <0.001 |

### Laboratory Parameters in Discharged and Died Patients

Median values of ALC were significantly lower (p < 0.001) in those who died (ALC < 1.0) versus those who were discharged (ALC > 1), during both waves. Median values of CRP, IL-6 and D-Dimer were three to four times higher in those who died, relative to those who survived in both the waves (P <0.001) but the values of LDH, Ferritin, CPK and Trop I were nearly two times (Table 6). This pattern was consistent in both the waves and the differences between the discharged and died were highly significant (P<0.001).

**Table 6.** Laboratory parameters in patients discharged and died in Wave-1 and Wave-2

| Laboratory parameter | Outcome | Wave-1 |  |  |  | Wave-2 |  |  |  |
| --- | --- | --- | --- | --- | --- | --- | --- | --- | --- |
|  |  | n | Median | IQR | P-value for comparison of discharged with deaths | n | Median | IQR | P-value for comparison of discharged with deaths |
| ALC ($10^9/L$) | Discharged | 11099 | 1.24 | 0.83-1.77 | <0.001 | 4030 | 1.08 | 0.74-1.54 | <0.001 |
|  | Death | 970 | 0.84 | 0.54-1.31 |  | 521 | 0.72 | 0.46-1.09 |  |
| CRP (mg/dl) | Discharged | 8910 | 7.7 | 1.7-27.3 | <0.001 | 3300 | 11.8 | 2.9-45.9 | <0.001 |
|  | Death | 685 | 26.5 | 9.6-116.5 |  | 352 | 50.3 | 13.5-122.3 |  |
| IL 6 (pg/ml) | Discharged | 7241 | 18.9 | 6.8-48.1 | <0.001 | 2613 | 18.6 | 6.8-44.9 | <0.001 |
|  | Death | 690 | 91.8 | 35.3-243.6 |  | 329 | 67.8 | 23.9-161.8 |  |
| D-Dimer (ng/ml) | Discharged | 8817 | 217 | 135-380 | <0.001 | 3223 | 220 | 150-357 | <0.001 |
|  | Death | 693 | 738 | 342-2417 |  | 362 | 536.5 | 292.3-1653 |  |
| LDH (U/L) | Discharged | 6474 | 288 | 229-375 | <0.001 | 2193 | 290 | 229-382 | <0.001 |
|  | Death | 538 | 523.1 | 380-733 |  | 290 | 499.4 | 360-765 |  |
| Ferritin (ng/ml) | Discharged | 7244 | 216.2 | 94.4-445.6 | <0.001 | 2671 | 219.9 | 98.6-56.2 | <0.001 |
|  | Death | 258 | 287.1 | 158.1-531.9 |  | 310 | 473.6 | 246.6-944.4 |  |
| CPK (U/L) | Discharged | 3909 | 94.8 | 57-179 | <0.001 | 1500 | 103 | 61-196 | <0.001 |
|  | Death | 358 | 150.5 | 65-323 |  | 196 | 196 | 75.5-516.5 |  |
| Trop I (ng/ml) | Discharged | 3516 | 0.01 | 0.01-0.01 | <0.001 | 1445 | 0.01 | 0.01-0.01 | <0.001 |
|  | Death | 270 | 0.03 | 0.01-0.21 |  | 270 | 0.03 | 0.01-0.01 |  |

**Table 7.** Discharges, died, and total cases with different CTSS categories in Wave-1 and Wave-2

| CT SS | Wave-1 |  |  | Wave-2 |  |  |
| --- | --- | --- | --- | --- | --- | --- |
|  | Discharged<br>n = 5910 | Died<br>n = 301 | Total<br>n = 6211 | Discharged<br>n = 2626 | Died<br>n = 193 | Total<br>n = 2819 |
| Mild (0-9) | 2607 (44.1%) | 40 (13.3%) | 2647 (42.6%) | 1134 (43.2%) | 24 (12.4%) | 1158 (41.1%) |
| Moderate (10-15) | 2065 (34.9%) | 69 (22.9%) | 2134 (34.4%) | 1096 (41.8%) | 64 (33.2%) | 1160 (41.1%) |
| Severe (16-25) | 1238 (21.0%) | 192 (63.8%) | 1430 (23.0%) | 396 (15.1%) | 105 (54.4%) | 501 (17.8%) |
| Mean (SD) | 10.7 (5.8) | 16.6 (5.7) | 10.9 (5.9) | 10.2 (5.2) | 15.8 (5.6) | 10.6 (5.4) |
| P-value for comparison with Wave-1 |  |  |  | <0.001 | 0.126 | 0.022 |

### CT Severity Score

CTSS was available for 6211 patients (43.1% of total patients) of Wave-1 and 2891 patients (53.0% of total patients) of Wave-2. Patients with moderate (10-15) value of CTSS were higher (41.1%) in Wave-2 than in Wave-1 (34.4%) and correspondingly lower with mild (0-9) and severe (16-25) values. In both the waves, mean value in those who died was nearly 50% higher than in those who survived (P <0.001 for both the waves). The ratio of discharged to died in the cases with severe values was nearly 1:6 in Wave-1 but increased to 1:4 in Wave-2.

## DISCUSSION

Several countries experienced second wave of COVID-19 pandemic. In most parts of the world, the number of people infected with COVID-19 are reported to be more in the second wave than in the first wave [2, 5]. India also witnessed a devastating second wave, which peaked in April-May 2021. Reasons for the explosive second wave in India would be multifactorial but it was arguably triggered by emerging lineage of SARS-COV-2 variants B.1.617, particularly its sub-lineage B.1.617.2, now called delta variant[8].Lack of COVID-appropriate behaviour (use of mask and social distancing) amongst people in the wake of vanishing first wave may have aggravated the incidence. Accumulating evidence suggests that B.1.617 lineage variants are more transmissible and perhaps more lethal than B.1.1.7 (alpha variant) [2, 4], which had been a dominant strain in Indian population before the arrival of second wave [8]. This could have caused a substantial shift in the profile, management, and outcome of the cases in the second wave.

We undertook this retrospective study to investigate differential characteristics of COVID-19 patients admitted in the second wave vis-à-vis the first wave. All cases admitted in 2020 were considered belonging to the first wave, designated as Wave-1, and the cases admitted in 2021 were considered to belonging to the second wave, designated as Wave-2. The study included 14398 cases of Wave-1 and 5454 cases of Wave-2. This might be the biggest series for comparison of the cases in the first and the second wave in India.

Higher percentage of patients with severe illness were admitted in Wave-2 compared to Wave-1 (39.4% vs 32.7%; P<0.001). Other than the agent factor (viral mutant B.1.617.2 which is probably more virulent), other factors for increased disease severity could be late presentation, as during the peak of Wave-2 due to extreme shortage of beds in wards and ICUs of most hospitals patients were being managed at home on oxygen. By the time they could get a bed in the hospital, they were already in moderate or severe hypoxemic failure. Moreover, in the early phase of pandemic in Wave-1, in India, according to the initial guidelines of the Government, all RT-PCR positive COVID-19 patients, irrespective of their disease severity, were admitted including mild cases in April-May 2020.

Epidemiological features of the second wave (August-September 2020) and the third wave (from November 2020) of COVID-19 in South Korea was compared and descriptive analysis was done [9]. The authors noted the delayed strengthening of social distancing polices in the third wave (15 days) compared to the second wave (3 days), and higher case fatality rate in the third wave (1.26% vs 0.9%), clearly indicating the importance of social distancing in the absence of effective vaccines to control the spread of SARS-CoV2 and unavailability of antiviral drugs to treat the cases. [9]

Age distribution of the cases, in our study, remained nearly the same in both the waves and older age group (>60 years) comprised almost 40% of the admitted patients. This happens to be nearly 4 times of their share in the general population and indicates their high vulnerability in both the waves. Study from India by Jain et al. [10] observed that, in addition to the older persons, the paediatric and younger individuals were also more infected in India in the second wave [10]. We did not observe any such pattern in our cases, and the belief in some quarters that the second wave affected more of the younger people is not borne out by the admitted cases in our hospitals. Iftimie et al. [11] conducted a prospective study in Spain and compared characteristics of the two waves in hospitalised patients using data from two equal periods of 3½ months. In their cohort, the number of patients admitted in the first and second wave was 204 and 264, respectively. They reported that hospitalised patient in the second wave were significantly younger with mean age of 57± 26 years vs. 67±18 yrs. in the first wave) (P<0.001). They also reported that in the second wave, children in age group (<15 years or what) were high, although vast majority of them had mild disease that did not require hospitalization for less than 4 days.

We found that a higher number of patients with comorbidities were admitted in Wave-2 compared to Wave-1 (59.7% vs. 54.8%; P<0.001). More than 40% of the admitted patients in both the waves had diabetes (DM) or hypertension (HTN) or both. Chronic kidney disease was reported in 13% and 15% of the patients, respectively, while CAD was in 5% and 6% of the cases. Significantly more patients with DM (P=0.031), HTN (P=0.001) and CKD (P=0.004) were admitted during Wave-2. Iftimie et al. [11] reported from Spain that frequency of concomitant disease in cases of both the waves in their study did not show any significant difference. A retrospective study comparing the epidemiology, clinical presentation and outcomes of case in the two waves of COVID-19 was done in patients from South Africa by Maslo et al. [12] and observed that patients hospitalised in the second wave were significantly older (median 57 yrs. vs. 54 yrs.) and had fewer co-morbidities than in the first wave. Kurriet et al. [13] conducted a hospital-based retrospective study in a total of 84 ICU patients, and nearly 78% of them had one or more underlying co-morbidities, hypertension followed by diabetes being the most common.

In our series, nearly one-third of patients during both the waves were admitted to ICU / HDU. However, the average duration of hospitalization and readmission rates were lower in Wave-2 compared to Wave-1. This might be a reflection of change in the discharge policy of the Ministry of Health and Family Welfare, Government of India, where, during most part of 2020 (Wave-1), the admitted patients needed to be tested negative twice by RT-PCR for COVID-19 before discharge and thus many patients needed to stay much longer, waiting to convert negative, in spite of being asymptomatic. This policy was revised, and a negative RT-PCR report was removed as a prerequisite for discharge, except in severely ill patients. Better standard operating procedures and availability of more therapeutic options during the latter phase of Wave-1 and during Wave-2, could be the other reasons contributing to reduced days of hospitalization in the second wave. Iftimie et al. [11] also observed for Spain that the duration of hospitalisation was significantly shorter in the second wave (14 ± 19 days vs. 22 ± 25 days).

We observed that more patients required oxygen support in Wave-2 (74.1% vs. 63.4% in Wave-1; P<0.001) and similarly a higher number required invasive ventilation (10.1% vs. 8.7% in Wave-1; P=0.002). Maslo et al. [12] reported for South Africa that ICU admission and use of mechanical ventilation was in lower proportion of cases in the second wave. The higher use in our cases could be again due to their late arrival in the hospital due to acute shortage of beds during the peak occurrence.

In our cohort of patients, remdesivir usage in Wave-2 was much higher than in Wave-1 (74.4% vs. 55.3%). This was expected as remdesivir only became available for use in India only after mid-May 2020. Usage of steroids was high but almost similar during the two waves (nearly 86%) and so also of anticoagulants like enoxaparin (74-77%). These two drugs had become the standard of care for most of the hospitalized patients with moderate to severe COVID-19 as per the guidelines of the Ministry of Health in May 2020. However, use of IVIG during Wave-2 was much higher than in Wave-1 (3.7% vs. 2.0%), while that of favipiramir and tocilizumab was reduced to almost half in Wave-2 compared to Wave-1. The use of convalescent plasma remained almost same in both the waves --it was given approval for an “Off-label” use in moderate and severe COVID-19 in June 2020 by the Drug Controller General of India [7]. Iftimie et al. [11] found in their study in Spain that cases in the second wave were more often treated with non-invasive ventilation and corticosteroids and less often with invasive mechanical ventilation, conventional oxygen therapy and anticoagulants. Maslo et al. [12] reported for South Africa that management of COVID-19 in the two waves was different as use of remedesvir and tocilizumab was more in the second wave than in the first, while use of corticosteroids and anticoagulant seen in around 90% patients of both the waves. A study from France [14] compared the characteristics and outcome between the patients admitted to COVID-19 ICU for acute respiratory failure during the first (March 13 – May 27, 2020) and second wave (August 19 – December 7, 2020) of COVID-19. They had 82 patients in the first wave and 50 during the second wave in ICU. During the second wave, patients admitted in ICU received early glucocorticoids and intermediate /full-dose thrombo-prophylaxis, lower proportion of cases required invasive mechanical ventilation and had lower rate of thrombotic event, in comparison to the first wave. However, overall ICU morality and duration of ICU stay did not differ in the two waves of COVID-19. [14]

As the overall disease severity of cases was higher in Wave-2, the trend of inflammatory blood markers also reflected the same. Median values of ALC were significantly lower and those of CRP, D-dimer, LDH and CPK were significantly higher in Wave-2 than in Wave-1. The high levels of these markers also correlated with higher mortality in Wave-2 cases. Analysis of laboratory parameters, in the study by Maslo et al. [12] indicated increased risk of severe COVID-19 in the second wave. D-dimer and IL-6 levels on admission were significantly higher in the second wave compared to the first wave (1.1 vs. 0.9 mg/l and 53.1 vs. 32.4 pg/ml, respectively) in their cases, indicating higher disease acuity on admission in the second wave. However, other markers of severity, i.e., NLR and CRP level were nearly the same in both the waves. Kurriet et al. [13] found that severe COVID-19 was associated with a high NLR and moderately elevated inflammatory markers (CRP, Ferritin, IL-6) and thus can be used for prognostication.

In our study, a high proportion of patients admitted in Wave-2 had secondary bacterial and fungal infection than in Wave-1 (27.6% vs. 11.8%) (P<0.001). UTI was the commonest secondary infection, followed by BSI, HAP and SSTI. The broad spectrum of microbes causing these infections included mostly Gram negative bacilli of Enterobacteriace group and Candida species. In the months of May and June 2021, there was a sudden spurt in COVID-19 patients being referred to our hospitals with secondary Mucormycosis (COVID-associated Mucormycosis (CAM). We reported 169 cases of CAM and 17 deaths in Wave-2 (in just about 6 to 8weeks period) while in Wave-1, the number was 10 with 2 deaths. These were associated with diabetes, uncontrolled blood sugars due to steroid use and high ferritin levels. A reterospective study by Vijay et al. [15] of secondary infection in COVID-19 cases admitted in 10 hospitals of India between June and August 2020 showed, that 3.6% cases developed secondary bacterial/fungal infection and of which 56.7% died, i.e. very high rate of mortality in cases with secondary infection. Predominance of Gram negative pathogens (78% cases) with high rates of carbapenem resistance was reported in their cohort, with Klebsiella pneumonia (29%) and Acinetobacter baumanii (21%) most common pathogen. [15]

The lower case fatality rate due to COVID-19 in Wave-2 has been reported in multiple studies across the globe [11, 16, 17], while few studies reported higher case fatality rate [12]. India, till June 21, has experienced two waves of COVID-19, first in 2020 and second in early 2021. Infection fatality rate (IFR) in these waves varied largely along with underreporting of COVID-19 infection and death in India. Purkayartha et al. [16] tried to reconcile the estimates of IFR arrived from sero-prevalence studies with epidemiological model based estimates, and also compared the estimated IFR of Wave-1 and Wave-2. They estimated underreporting factor for infection to be 11.11 and for death 3.56 for Wave-1. While in Wave-2, underreporting factor escalated to 26.77 for infection and 5.77 for death. Taking these factors into account, the IFR estimate was 0.46% for the first and 0.18% for the second (till May 15). [16]

We found an overall mortality in Wave-2 nearly 40% higher than in Wave-1 (10.5% vs. 7.2%; P<0.001). Patients less than 40 years recorded the sharpest increase in mortality in Wave-2 (4.1% vs. 1.3%), with higher mortality in males. In all other age groups also, the mortality in Wave-2 was significantly higher than in Wave-1. A higher mortality in Wave-2 was also observed in patients irrespective of their ventilator requirement and those who received and not received convalescent plasma. Not only was the mortality higher in Wave-2 for patients in ICU (19.8% vs. 25.1%) but steeply higher for those admitted in ward (0.5% vs. 3.1%). This was mostly because many sicker patients on high flow oxygen had to be treated in wards as there were no ICU beds available, during the peak of Wave-2. Our analysis of the duration of symptoms at admission in young patients (<45 years of age) who died in both the waves revealed that on average, the patients in second wave came a day late (6.3 days vs 7.3 days). An analysis of the data from 14 countries showed age distribution of COVID-19 deaths to be fairly similar in the second as in the first wave [18]. Study from India by Jain et al. reported that the total number of deaths during second wave of COVID-19 was high due to alarmingly high number of cases, but the death rate showed no significant increase [10]. However, the results of study by Iftimie et al. [11] from Spain showed that the case fatality rate decreased from 24% in the first wave to 13.2 % in the second wave. Those died in the second wave were much older than those died in the first wave. Graichen [19] described the difference between the first and second wave from a European-German prospective. The number of deaths in Germany due to COVID-19 in a period between March-October 2020 summed up around 9000, while in 10-12 weeks of second wave more than 20,000 people were reported dead. The increase in mortality in the second wave was noted in many countries, despite improved management protocols due to the knowledge gained from the first wave. In the study by Maslo et al. [12] the overall mortality due to COVID-19 was reported around 32.6% and 36.4% in the first wave and second wave in South Africa respectively. However, ICU mortality was significantly high in the second wave compared to the first wave (74.4% vs. 57.1%). Considering that the patients in the second wave in that country were infected with 501Y.V2 variant of SARS-CoV2, increased fatality rate indicates the increased virulence of this variant. [12]

In our study, the average CT severity score (CTSS), in both the waves was nearly 50% higher in those who died compared to those who survived (P<0.001). Otherwise, the percentage of patients with moderate score (10 -15) was higher in Wave-2 compared to Wave-1 41.0% vs. 34.0%), but this was not with the cases with severe CTSS of more than 15 (23% in Wave-1 vs 18% in Wave-2). One reason for this could be that this diagnostic modality might have been used earlier in the course of illness. Kurriet et al. [13] reported in their study that the average CTSS was 12.43 ± 5.7 in survivors, while CTSS for deceased was 18.87 ± 4.68 (P<0.0001). ICU non-survivors were found to be more hypoxic on admission (SpO2 = 75.07) when compared with the survivors (SpO2 = 81.96). Strong positive correlation was reported between the Fio2 at admission and CTSS; and between D-dimer and duration of oxygen requirement. Kurriet et al. [13] reported the disease severity significantly correlated with CTSS and D-dimer, and CTSS >15 and D-dimer >2.4 correlated strongly with mortality. With the increasing experience and knowledge of disease management, the protocols for management included the inflammatory markers and CT scan of chest to assess disease severity.

## CONCLUSIONS

Overall higher severity of the disease at admission and a significantly a higher mortality rates in the second wave, especially in younger patients, were the main findings of the study. Since there were no significant demographic differences in the population during these two waves, various other factors such as increased comorbidity and higher occurrence of secondary bacterial and fungal infections may have contributed towards increased mortality. Reports indicate higher percentage of infections having been caused by delta variant (B.1.617.2) in the second wave, which was not only more transmissible but also potentially more lethal, could be another important factor. Late presentation of patients in wave 2 due to non-availability of hospital beds could also have contributed towards higher mortality. All these factors need further studies for delineating exact role.

## Data Availability

All the admitted patients data taken from EHR.

## ROLE OF AUTHORS

SB designed the study concept and contributed patients for the study, AI did the statistical analysis, MA wrote the manuscript, VJ and DJ did the data access and collection from EHR, BT provided microbiological data for secondary infection, PD provided lab data of COVID patients, BA provided CTSS data for COVID patients, rest all are clinicians and contributed and treated patients in the present study.

## ACKNOWLEDGEMENTS

We wish to acknowledge and thank the contribution made by Taruna Sharma

